# Structural neuroplastic responses preserve functional connectivity and neurobehavioural outcomes in children born without corpus callosum

**DOI:** 10.1101/2020.05.29.20115451

**Authors:** Vanessa Siffredi, Maria G. Preti, Valeria Kebets, Silvia Obertino, Richard J. Leventer, Alissandra McIlroy, Amanda G. Wood, Vicki Anderson, Megan M. Spencer-Smith, Dimitri Van De Ville

**Affiliations:** Institute of Bioengineering, Center for Neuroprosthetics, Ecole Polytechnique Fédérale de Lausanne, Switzerland; Department of Radiology and Medical Informatics, Faculty of Medicine, University of Geneva, Switzerland; Brain and Mind Research, Clinical Sciences, Murdoch Children’s Research Institute, Melbourne, Australia; Division of Development and Growth, Department of Paediatrics, Faculty of Medicine, University of Geneva, Switzerland; Department of Electrical and Computer Engineering, Clinical Imaging Research Centre, N.1 Institute for Health and Memory Networks Program, National University of Singapore, Singapore; Department of Paediatrics, University of Melbourne, Melbourne, Australia; Department of Neurology, Royal Children’s Hospital, Melbourne, Australia; Neuroscience Research, Clinical Sciences, Murdoch Children’s Research Institute, Melbourne, Australia; School of Life and Health Sciences & Aston Neuroscience Institute, Aston University, Birmingham, B4 7ET UK; School of Psychology, Faculty of Health, Melbourne Burwood Campus, Deakin University, Geelong, Victoria, Australia; School of Psychological Sciences, University of Melbourne, Melbourne, Australia; Department of Psychology, Royal Children’s Hospital, Melbourne, Australia; Turner Institute for Brain and Mental Health, School of Psychological Sciences, Monash University, Melbourne, Australia

**Keywords:** callosal agenesis, brain plasticity, structural connectivity, functional connectivity, structural reorganisation

## Abstract

**Background:** The corpus callosum is the largest white matter pathway in the brain connecting the left and the right hemispheres. Developmental absence of the corpus callosum is a model disease for exploring disrupted connectivity and in turn understanding plasticity of the human brain, with atypically developing structure and function resulting in a highly heterogeneous clinical and cognitive profile. A proposed candidate for neuroplastic response in the context of this brain malformation is strengthening of intra-hemispheric pathways.

**Methods:** To test this hypothesis, we assessed structural and functional connectivity at the whole-brain and regional level in a uniquely large cohort of children with agenesis of the corpus callosum (AgCC, n = 20) compared with typically developing controls (TDC, n = 29), and then examined associations with neurobehavioural outcomes using a multivariate data-driven approach.

**Results:** For structural connectivity, children with AgCC showed a significant increase in intrahemispheric connectivity in addition to a significant decrease in inter-hemispheric connectivity compared with TDC. In contrast, for functional connectivity, children with AgCC and TDC showed a similar pattern of intra-hemispheric and inter-hemispheric connectivity. In AgCC, structural strengthening of the intra-hemispheric pathway was uniquely associated with verbal learning and memory, attention and executive measures.

**Conclusions:** We observed structural strengthening of intra-hemispheric pathways in children born without corpus callosum, which seems to allow for functional connectivity comparable to a typically developing brain, and were relevant to explain neurobehavioural outcomes in this population. This neuroplasticity might be relevant to other disorders of axonal guidance, and developmental disorders in which corpus callosum alteration is observed.

## INTRODUCTION

Neuroplasticity is the intrinsic property of the central nervous system to respond dynamically to the environment via modification of neural circuitry. In the context of a brain lesion, plasticity provides a mechanism for the brain to adjust through remyelination, reorganisation of circuits, and/or neural and behavioural compensation (1, 2). In particular, clinicopathological observations suggest that the immature human brain is capable of major structural and functional reorganisation (3). As an example of early disruption of programmed developmental brain processes, children and adults born without a corpus callosum show remarkable brain plasticity. Indeed, there is very little evidence of the interhemispheric disconnection compared to adults who have undergone surgical disconnection of the corpus callosum (i.e., split brain patients) which affects motor control, spatial orientation, vision, hearing, and language (4).

With more than 190 million topographically organised axons, the corpus callosum is the major commissural fibre bundle in the human brain connecting homologous structures in both hemispheres (5). This structure develops prenatally with the first fibres crossing the midline through the massa commissuralis as early as 11–12 weeks of gestation (6, 7), and reaches its final shape by 20 gestational weeks. In typical brain development, these callosal connections participate in a myriad of lower and higher level cognitive functions, and in the integration of complex information transfer between the cerebral hemispheres (8). Given the structural and functional importance of the corpus callosum and its very early maturation, its developmental absence, called agenesis of the corpus callosum (AgCC), provides a unique window to better understand neuroplastic responses.

In the context of AgCC, different neuroplastic responses, for example, enlarged (hyperplasia) of anterior and posterior commissures, may contribute to neurobehavioural functioning (9, 10). A recent study found associations between structural and microstructural properties of these commissures and attentional capacity (10). Moreover, atypical pathways connecting the posterior parietal cortices homotopically via the anterior and the posterior commissures, as well as the primary visual areas via the anterior commissure, have been reported in case studies (11, 12). Another proposed neuroplastic response is a functional compensation via strengthening of intrahemispheric pathways in individuals with AgCC (13–15). In foetuses with AgCC (n = 20), Jakab and colleagues (2015) showed weaker inter-hemispheric structural connectivity starting in the second trimester of gestation followed by stronger intra-hemispheric structural connectivity starting specifically in the third trimester, compared to control foetuses with normal brain development. To date, strengthening of intra-hemispheric pathways has not been explored in individuals born with AgCC, and the potential implications of this neuroplastic response for neurobehavioural outcomes are unknown.

AgCC is a heterogeneous condition. It can be complete or partial, and may be isolated or associated with additional brain abnormalities such as cortical malformations (5, 17). Individuals with AgCC present with impairments of varying severity across a range of neurobehavioural domains (17, 18). General cognitive ability in individuals with AgCC who come to clinical attention is typically in the low average range (18, 19), with difficulties in executive functions commonly observed, including deficits in cognitive control such as selective attention and inhibition, as well as cognitive flexibility (19–21). Learning and memory difficulties (22, 23), cognitive processing speed (20, 22), and difficulties in social and emotional cognition (24) may also be present. Consistent with the heterogeneity of their neuroimaging profile, neurobehavioural outcomes vary (19), and the role of neuroplastic responses in explaining this variability remains unclear.

This study aimed to assess brain structural and functional connectivity at the whole-brain and regional level, and their associations with neurobehavioural outcomes across general cognitive, executive, memory and social domains, in a uniquely large cohort of children with AgCC compared with typically developing controls (TDC). We hypothesised a neuroplastic response resulting in strengthening of intra-hemispheric pathways in children with AgCC accounts for the documented variability in neurobehavioural outcomes in this population.

## METHODS AND MATERIALS

### Sample

This study used data from the “Paediatric Agenesis of the Corpus Callosum Project” (19), which examined neurobehavioural, neurological and neuroimaging outcomes in children with AgCC (n = 28) compared with typically developing children (n = 30). This project was approved by the Royal Children’s Hospital (RCH) Human Ethics in Research Committee. Written informed consent was obtained from the parents or legal guardians prior to participation. A cohort of 28 children with AgCC was recruited. Inclusion criteria were: 1) aged 8 years 0 months to 16 years and 11 months; 2) documented evidence of AgCC on Magnetic Resonance Imaging (MRI) conducted as part of a routine clinical work-up; 3) English speaking; and 4) functional ability to engage in the assessment procedure. MRI findings were qualitatively reviewed by a paediatric neurologist with expertise in brain malformations (RJL), who confirmed diagnosis of AgCC, including complete and partial AgCC, and identified associated brain malformations. A TDC group of 30 children comparable in age and sex to the AgCC group was recruited through advertisement in local schools and through staff at the RCH.

### Procedure

Consenting families were seen at a research clinic at the Murdoch Children’s Research Institute. Child neurobehavioural assessment was conducted by a training psychologist with expertise in assessments in children, using standardised tests that were developmentally appropriate for children and commonly used in clinical settings.

## Materials

### Neuroimaging

#### Image acquisition

Images were acquired on a 3T MAGNETOM Trio scanner (Siemens, Erlangen, Germany) at the RCH. A 32-channel head coil was used for transmission and reception of radio-frequency and signals. An anatomical image was acquired using a T1-weighted MP-RAGE sequence (TR = 1900 ms, TE = 2.71 ms, TI = 900 ms, FA = 9°, FoV = 256mm, voxel size = 0.7 × 0.7 × 0.7 mm). Echo planar diffusion-weighted imaging (DWI) data were acquired at two different b-values, including two scans without diffusion weighting (b-factor = 0): a) b-value = 3000s/mm2, 50 gradient directions where 54 slices with isotropic voxels of 2.3 mm3 were obtained (TR = 8200 ms; TE = 112 ms; FoV = 240 mm) in an anterior to posterior direction; and b) b-value = 1000s/mm2, 30 gradient directions where 64 slices with isotropic voxels of 2 mm3 were obtained (TR = 8600 ms; TE = 90 ms; FoV = 256 mm) in an anterior to posterior direction. Resting-state gradient-echo EPI sequences was also acquired (196 frames, TR = 2000ms, TE = 30ms, voxel size: 2.6 × 2.6 × 4 mm, FA: 90 deg, FoV = 250 mm × 250 mm). During the resting-state functional MRI (fMRI) sequence, participants were instructed to keep their eyes closed and engage into mind wandering.

#### Structural and functional MRI data preprocessing and analyses

A flowchart summarises brain connectivity preprocessing and analyses, see Figure 1.

**Figure 1.**
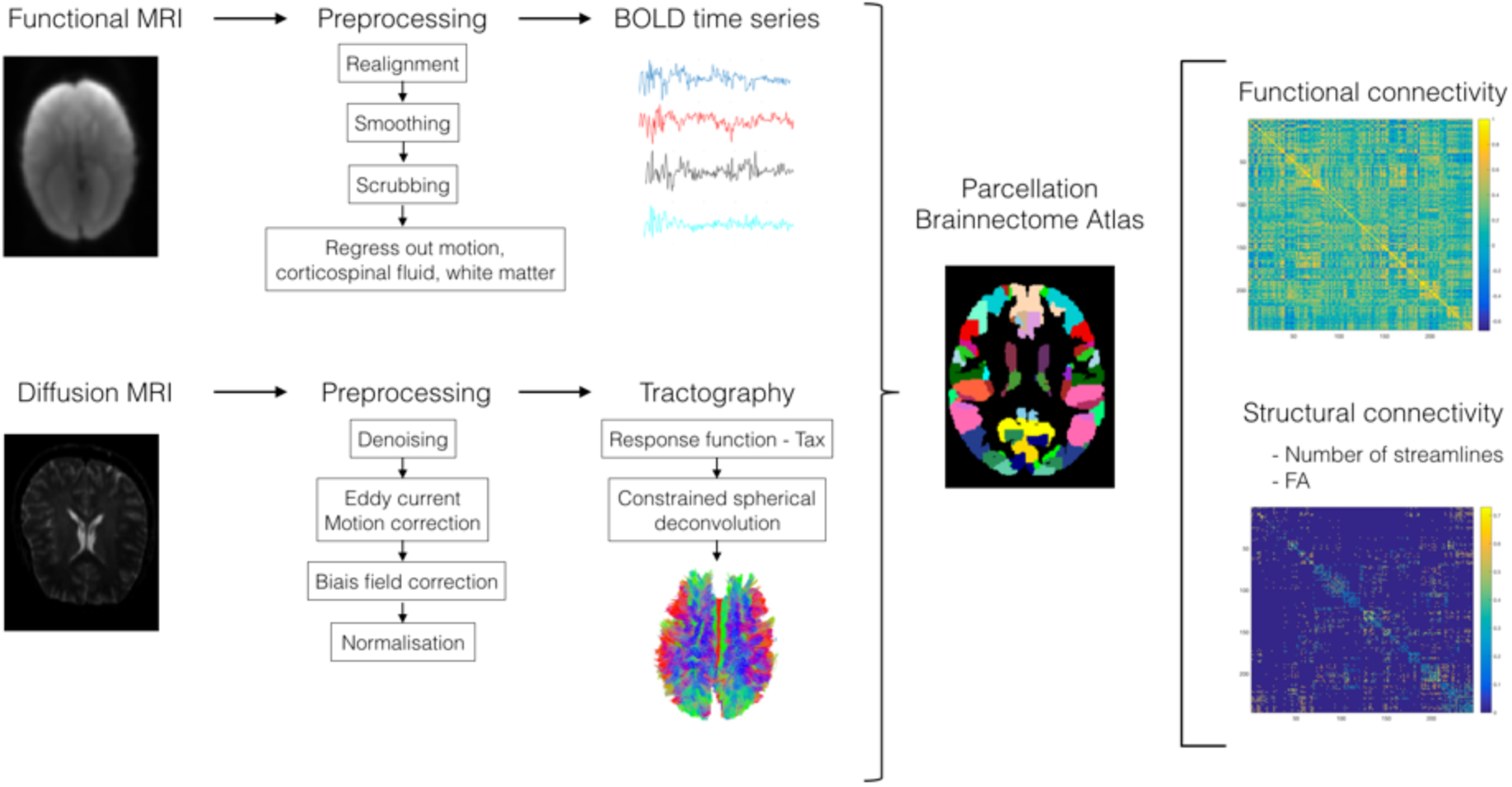
Flowchart summarising the computation and analysis of functional and structural brain connectivity. Briefly, fMRI BOLD time courses / DWI fibre pathways were extracted, both data were parcellated using the Brainnetome atlas. Individual connectivity matrices were generated by computing pairwise functional connectivity (i.e., Pearson’s correlations) between regional timeseries and structural connectivity (i.e., number of streamlines and mean FA) between brain regions.

Briefly, DW images were preprocessed using MRtrix 3 (25). Probabilistic tractograms and subsequent filtering using Spherical Deconvolution Informed Filtering of Tractograms were completed (26). For each participant, the structural connectome matrix was generated from the resulting tractography using the registered Brainnetome Atlas (27). Two 246 × 246 connectivity matrices, referred as SC, were obtained using the following connectivity metrics: a) number of streamlines weighted for sum of the volumes of the connected regions; and b) average fractional anisotropy (FA).

Resting-state fMRI data were preprocessed and analysed using SPM12 and the pipeline described by Preti and Van De Ville (2017). We implemented volume censoring (‘scrubbing’) for motion correction using a framewise displacement of 0.5 mm threshold for exclusion (28, 29). Region-averaged time series were extracted in each participant’s individual space using the registered Brainnetome Atlas. We computed pairwise Pearson correlation coefficients between all pairs of regions to obtain a symmetric correlation matrix of 246 × 246 for each participant, referred to as functional connectivity (FC).

For both SC and FC measures, average values of all inter-hemispheric connections (left-right), and intra-hemispheric connections (right to right and left to left) were computed for each participant. Lobe-wise SC and FC was also computed for intra- and inter-hemispheric connectivity, see Supplemental Methods and Materials.

### Neurobehavioural outcome measures

Associations with SC and FC and variation in neurobehavioural outcomes were examined using 33 age-standardised measures administered as part of the “Paediatric Agenesis of the Corpus Callsoum Project” assessing five neurobehavioural domains; general cognitive functioning, short-term and working memory, executive and attentional functions, learning and memory, as well as social functions, see Supplemental Methods and Materials and supplementary Table S1.

### Statistical analyses

Group differences in whole brain intra-hemispheric and inter-hemispheric connectivity for SC (number of streamlines and mean FA) and FC were compared between the following groups: a) children with AgCC versus TDC; b) complete versus partial AgCC; and c) isolated versus complex AgCC (i.e., AgCC associated with other brain malformations). Group differences between children with AgCC and TDC were also examined for lobe-wise intra- and inter-hemispheric SC and FC. When exploring data distributions in the two groups, none of the assumptions for parametric tests were respected. Therefore, group differences were investigated with the Wilcoxon Signed-Ranks Test using R software Version 1.1.463 (http://www.r-project.org; R Project for Statistical Computing, Vienna, Austria). P values, as well as q values corrected for multiple comparisons using a False Discovery Rate (FDR, q<0.05) correction (30), were reported for all SC and FC comparisons (n = 75).

To evaluate associations between connectivity (i.e., intra- and inter-hemispheric SC and FC) and neurobehavioural measures, we used Partial Least Squares Correlation (PLSC) analyses. PLSC is a multivariate data-driven statistical technique that maximises the covariance between two matrices by deriving latent components, which are optimal linear combinations of the original matrices (31) and described in previous work (32, 33). We used an in-house Matlab code to perform the analysis: https://miplab.epfl.ch/index.php/software/PLS (32, 34), see Supplemental Methods and Materials. Two separate PLSC analyses were conducted to test for associations between neurobehavioural variables and SC, and between neurobehavioural variables and FC, respectively.

## RESULTS

### Sample characteristics

Children were included in the current study if they had completed the required MRI sequences (T1, DWI and resting-state fMRI), resulting in the exclusion of five participants from the AgCC group and one participant from the TDC group. After quality checking the functional and structural MR images, three AgCC participants were excluded from the structural connectivity analysis, while six AgCC and one TDC participants were excluded from the functional connectivity analysis. Seven children were assessed on two separate occasions, for whom the most complete of the two neurobehavioural assessments was used, as well as the MRI scan complete at the time of the most complete neurobehavioural assessment.

The characteristics of the participants in the AgCC and the TDC groups are presented in Table 1 for each of the SC and FC analyses, given the samples differed for the two analyses. Twenty children with AgCC, including 13 with complete AgCC (4 isolated, 9 complex) and seven with partial AgCC (3 isolated, 4 complex), as well as 29 TDC, were included in the SC analysis. For the FC analysis, 16 children with AgCC, including 10 with complete AgCC (3 isolated, 7 complex) and six with partial AgCC (3 isolated, 3 complex), as well as 28 TDC met the inclusion criteria.

**Table 1.** Characteristics of the agenesis of the corpus callosum (AgCC) and typically developing control (TDC) groups included in the Structural Connectivity and Functional Connectivity analyses.

|  | AgCC | TDC | Group comparison |
| --- | --- | --- | --- |
| **Structural Connectivity (SC)** |  |  |  |
| **n** | 20 | 29 | - |
| **Age in years, mean (SD)[range]** | 12.08 (2.63)[8.67-17.08] | 11.75 (2.32)[8-16.42] | t(47)=0.048, p=0.642 |
| **Sex, n** | 7 females, 13 males | 13 females, 16 males | $\chi^2(1, n=49)=0.473, p=0.491$ |
| **Handedness, n** | 11 R, 8 L, 1 M | 26 R, 3 L | - |
| **Full-Scale IQ, median [range]** | 75.5 [66-126] | 114 | W=964.5, p<0.001 |
| **Functional Connectivity (FC)** |  |  |  |
| **n** | 16 | 28 | - |
| **Age in years, mean (SD)[range]** | 12.16 (2.75)[8.67-17.08] | 11.71 (2.35)[8-16.42] | t(42)=0.567, p=0.572 |
| **Sex, n** | 6 females, 10 males | 12 females, 16 males | $\chi^2(1, n=44)=0.121, p=0.728$ |
| **Handedness, n** | 10 right, 5 left, 1 Mix | 25 Right, 3 Left | - |
| **Full-Scale IQ, median [range]** | 76 [67-126] | 114.5 | W=808.5, p<0.001 |

Note: Full-Scale IQ was measured using the Wechsler Abbreviated Intelligence Scale (WASI) or the Wechsler Intelligence Scale for Children, 4th edition (WISC-IV; n=3) where the mean standardised score is M=100 and SD=15; Handedness was estimated by the Edinburgh Handedness Inventory with the following scores: Right-handed=+40 to +100, left-handed=-40 to -100, mixed handed=-40 to +40.

Of note, there was a large variability in the Full-Scale Intellectual Quotient (IQ) score, ranging from 66 to 126 in children with AgCC, and three children had a FSIQ below 70 (i.e., 2 SD or more below the test mean, in the Extremely Low range). A sensitivity analysis excluding these three children from the FC and SC analyses did not change our results.

### Structural Connectivity

For a summary of the whole brain and lobe-based SC and FC findings, see Figure 1.

For whole-brain SC measures (number of streamlines and FA), inter-hemispheric connectivity was significantly reduced in children with AgCC compared with the TDC group (q<0.0001), see supplementary Figure S1 and Table S2. In contrast, whole-brain intra-hemispheric SC was significantly increased in children with AgCC compared with the TDC group (q<0.0001 to q = 0.006). Within the AgCC group, there were no significant differences in either intra- or inter-hemispheric whole-brain SC in children with complete compared with partial AgCC, or in children with isolated AgCC compared with complex AgCC, see supplementary Table S3 and S4.

At the lobe level, children with AgCC had significantly reduced inter-hemispheric connectivity (q≤0.0001 to q = 0.0339) and intra-hemispheric connectivity for all lobes (q≤0.0001 to q = 0.0201) compared with the TDC group, except for the number of inter-hemispheric streamlines connecting temporal lobes to all contralateral lobes and the mean FA of streamlines connecting the left temporal lobe to all contralateral lobes, see supplementary Table S5. Moreover, children with AgCC had an overall increase in the number of streamlines in the left occipital lobe compared to the TDC group. More specifically, children with AgCC showed a significant increase in the number of intra-hemispheric streamlines in the left hemisphere and in the number of streamlines connecting the left occipital lobe to all contralateral lobes. Finally, children with AgCC had reduced mean FA in all intra- and inter-hemispheric streamlines compared to the TDC group (q≤0.001 to q = 0.01).

### Functional Connectivity

For whole-brain FC, intra- and inter-hemispheric connectivity did not differ significantly between the AgCC and TDC groups. Within the AgCC group, there were no significant differences in either intra-hemispheric or inter-hemispheric whole-brain FC in children with complete compared with partial AgCC, or between children with isolated compared with complex AgCC, see supplementary Figure S2 and Table S6, S7, S8.

For all lobes considered (frontal, parietal, temporal, occipital), there was no difference in either intra- or inter-hemispheric FC between the AgCC and TDC groups, see supplementary Table S9.

### Associations between brain connectivity and neurobehavioural outcomes

For neurobehavioural scores and group comparisons see supplementary Table S10.

The PLSC analysis applied on SC and neurobehavioural measures in the AgCC and TDC groups identified three statistically significant CorrComps: CorrComp 1 (p = 0.002) explained 35% of the covariance between structural connectivity and neurobehavioural measures, while CorrComp 2 (p = 0.032) and CorrComp 3 (p = 0.020) explained 22% and 5%, respectively. Based on the amount of explained covariance and clinical interpretability of the results (34), we chose to focus on CorrComp1. CorrComp1 revealed common as well as distinct patterns of associations in each group, see Figure 2. Both the AgCC and TDC groups showed increased intrahemispheric and reduced inter-hemispheric mean FA and number of streamlines associated with better dual task performance, indexing multimodal divided attention and working memory. In the AgCC group, increased intra-hemispheric and reduced inter-hemispheric mean FA and number of streamlines were associated with better neurobehavioural outcomes, including verbal learning and memory (including working memory), as well as attentional and executive functions, specifically sustained, selective and divided attention, processing speed, switching, verbal fluency and inhibition. In contrast, in the TDC group, increased inter-hemispheric and reduced intra-hemispheric mean FA and number of streamlines were associated with better performance in similar neurobehavioural measures, including verbal learning and memory, as well as attentional and executive functioning such as processing speed, switching and verbal fluency, but also in general neurobehavioural functioning (FSIQ and PIQ).

**Figure 2.**
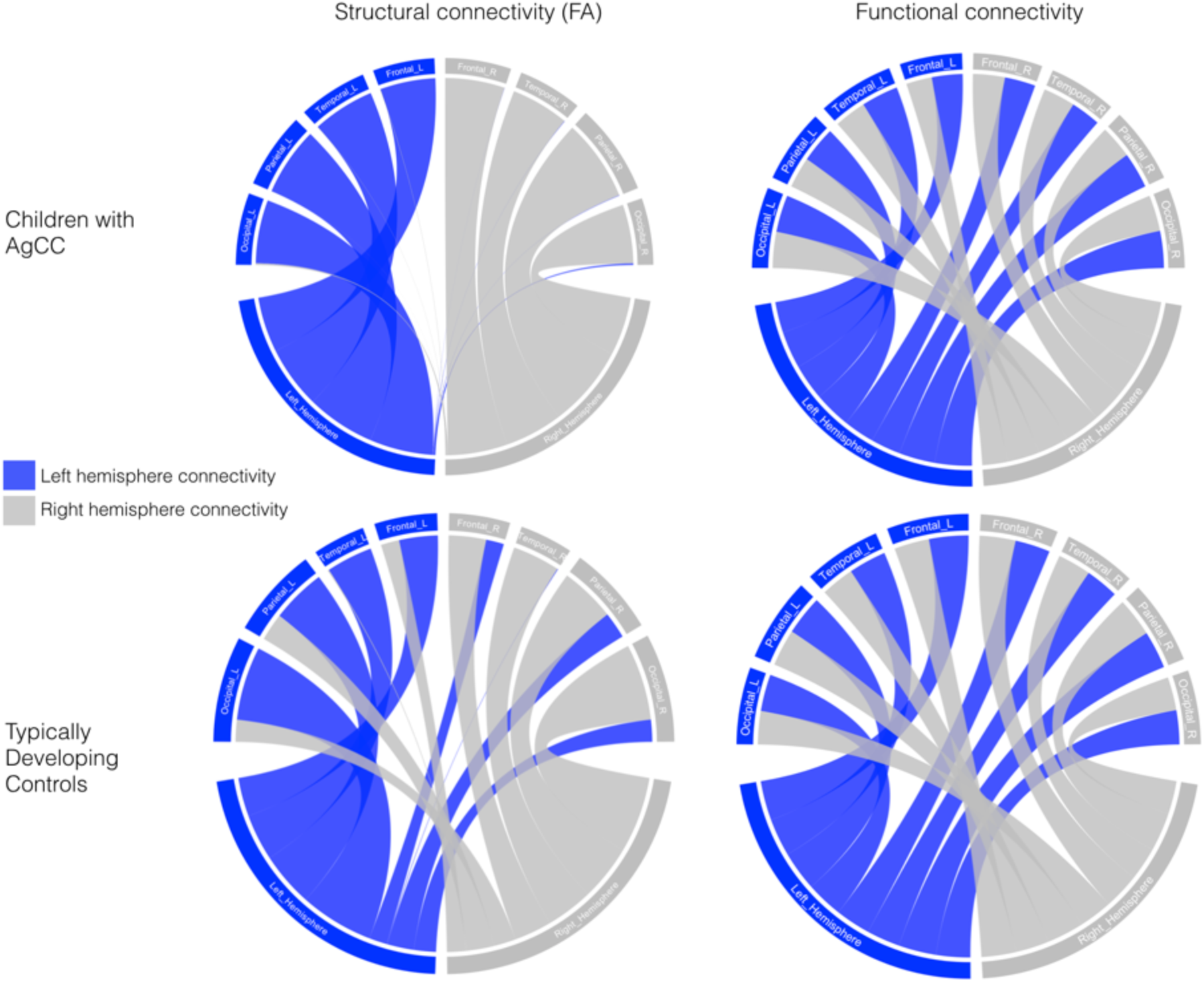
Summary of structural connectivity (fractional anisotropy, FA) and functional connectivity in children with AgCC and TDC. Grey lines represent connectivity in the right hemisphere, and blue lines represent connectivity in the left hemisphere. There is a shift from inter-hemispheric to intra-hemispheric structural connectivity in children with AgCC compared to the TDC group, while the pattern of functional connectivity is similar between the two groups.

The second PLSC analysis was applied on FC and neurobehavioural measures, and identified only one significant CorrComp (p = 0.002), which explained 96% of the covariance between FC and neurobehavioural measures. In both groups, increased intra-hemispheric and inter-hemispheric FC were associated with specific aspects of executive and attentional functioning, including visual scanning, selective attention and processing speed, see Figure 3. In the AgCC group, a robust negative association between FC and verbal recall was found. Interestingly, the association with dual task performance had an opposite effect in the two groups, with higher scores associated with an overall increase in FC in the AgCC group, whereas higher scores in the TDC group were associated with a decrease in FC. Finally, in the TDC group, a general increase in FC was also associated with VIQ.

**Figure 3.**
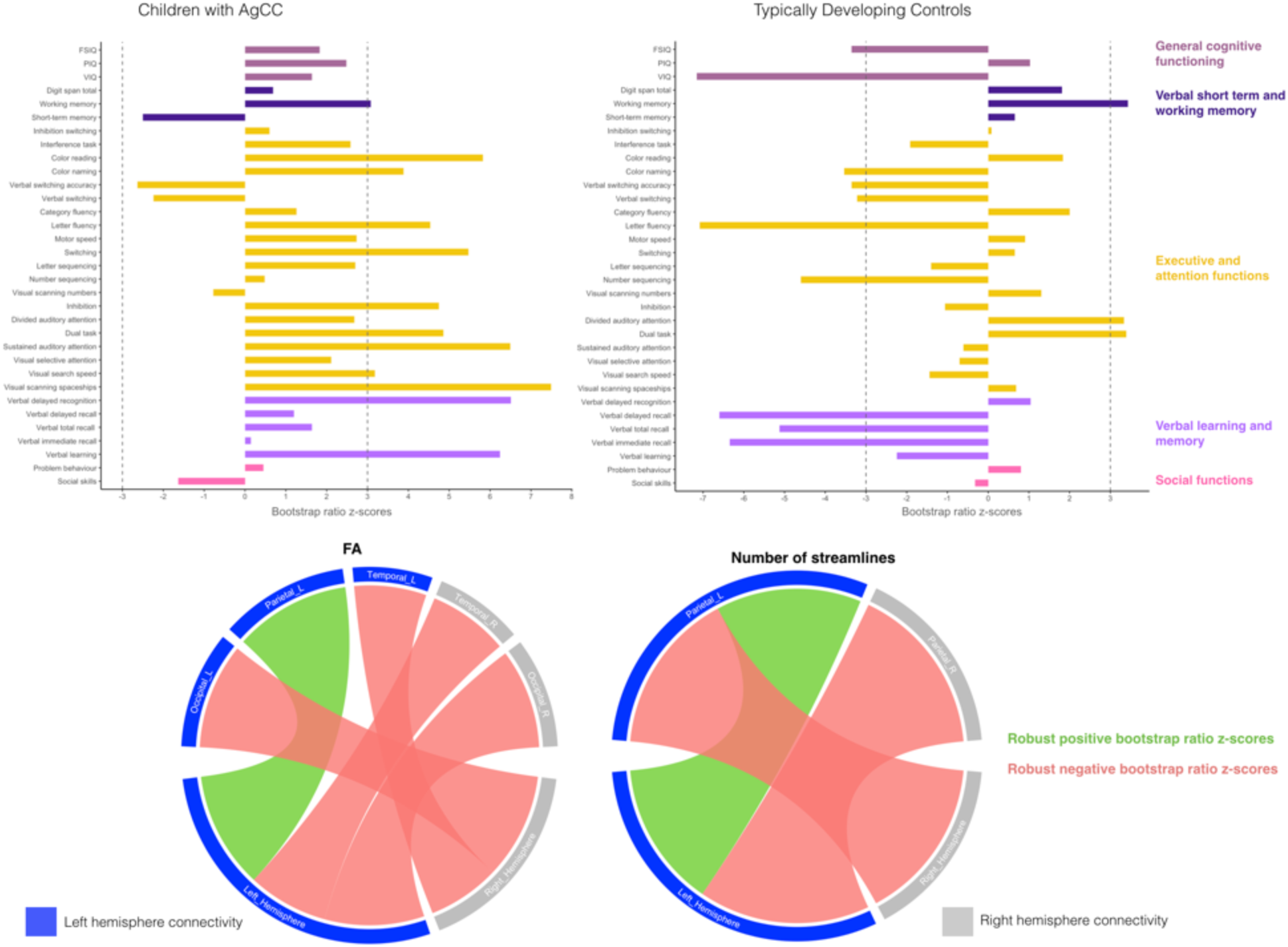
Associations between neurobehavioural outcomes and structural connectivity in the AgCC and TDC groups based on the PLSC analysis. Neurobehavioural weights are specific to each group, while brain connectivity weights are common to both the AgCC and TDC groups. Diverging graphs show bootstrap ratio z-scores (x axis) for each neurobehavioural measure (y axis); neurobehavioural measures are coloured by domains. Neurobehavioural measures with an absolute bootstrap ratio z-score above or equal to 3, indicated by a dash-dotted line on the graph, yield a robust contribution to the component. Circular graphs show robust bootstrap ratio z-scores (absolute bootstrap ratio z-scores above or equal to 3) for FA and number of streamlines lobe-wise in inter- and intra-hemispheric connectivity. Original saliences, as well as their bootstrap-estimated standard deviations and bootstrap ratio z-scores for all neurobehavioural and connectivity measures are reported in supplementary Table S11.

**Figure 4.**
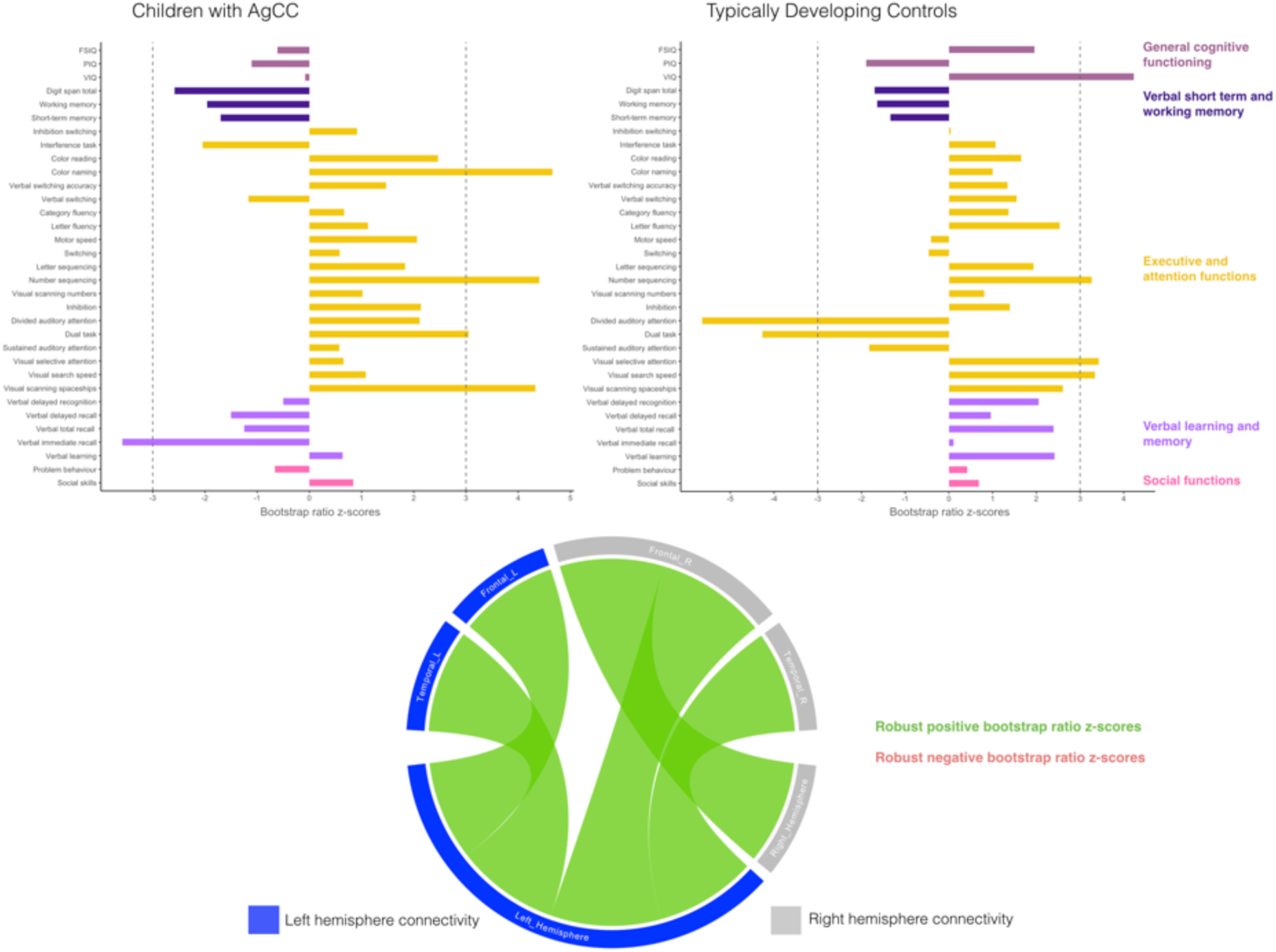
Associations between neurobehavioural outcomes and functional connectivity in the AgCC and TDC groups based on PLSC analyses. Neurobehavioural weights are specific to each group, while brain connectivity weights are common to both the AgCC and TDC groups. Diverging graphs show bootstrap ratio z-scores (x axis) for each neurobehavioural measure (y axis). Bootstrap ratio z-scores above or equal to 3, specify by a dash-dotted line on the graph, indicate a robust positive correlation; bootstrap ratio z-scores below or equal to –3, specify by a dash-dotted line on the graph, indicate a robust negative correlation. Circular graphs show robust bootstrap ratio z-scores for FC lobe-wise in inter- and intra-hemispheric connectivity; left regions and left hemisphere are in dark blue, right regions and hemisphere are in grey, positive robust correlations are indicated in green and negative robust correlations are indicated in orange at p<0.01. Salience, bootstrap-estimated standard deviations and bootstrap ratio z-scores for all neurobehavioural and connectivity measures are reported in supplementary Table S12.

## DISCUSSION

This is the first study to explore neuroplastic responses in a uniquely large cohort of children born without a corpus callosum using both structural and functional brain connectivity, and to examine their association with neurobehavioural outcomes. We show evidence of structural strengthening of intra-hemispheric pathways in children with AgCC, which seems to allow for inter- and intra-hemispheric functional connectivity comparable to typically developing brain. In children with AgCC, this increase in intra-hemispheric SC was associated with better neurobehavioural outcomes including verbal learning and long-term memory, verbal working memory, as well as executive and attentional functioning.

While whole-brain inter-hemispheric SC was reduced in children with AgCC compared to TDC, a significant increase in whole-brain intra-hemispheric SC was demonstrated in children with AgCC. These findings are in accordance with SC results of a previous study conducted in AgCC foetuses (16), suggesting that the neuroplastic responses began in utero and not in response to postnatal environmental factors. The neuroplastic responses observed could be linked to microstructural alterations that are known to take place at the cellular or synaptic level (35). It is possible that early in utero disruption in cortical developmental mechanisms allows for long-distance plasticity through atypical pivotal cellular and molecular mechanisms that divert axon growth and guidance through entirely new pathways and connections (11, 36). Regarding FC, and in contrast to the reduced inter-hemispheric FC reported in callosotomy patients (37), we found no difference between children with AgCC and TDC children for either intra-hemispheric or inter-hemispheric FC. Altogether, our findings are in line with the hypothesis of strengthening of intra-hemispheric pathways at the structural level as a neuroplastic response in children with AgCC, with a shift from inter-hemispheric connectivity to intra-hemispheric connectivity. They also imply that the relationship between SC and FC is regulated by complex and complementary processes (38). Structural neuroplastic responses through strengthening of intra-hemispheric pathways might underlie the compensatory mechanisms that lead to comparable patterns of intra-hemispheric and inter-hemispheric FC that is observed in a typically developing brain.

Complete and partial AgCC showed no difference in intra-hemispheric and inter-hemispheric SC or FC. As the corpus callosum develops very early during brain development, the immature human brain might engage similar neuroplastic responses at the whole brain level, independent of extent of the corpus callosum agenesis. In addition, children with isolated AgCC and complex AgCC (i.e., associated with other brain malformations) did not show any difference in either whole-brain intra-hemispheric or inter-hemispheric SC and FC. Therefore, despite the presence of additional brain malformations, children with complex AgCC seem to have a comparable level of plasticity in term of strengthening of intra-hemispheric pathways to those with isolated AgCC.

When exploring connectivity in different brain lobes, patterns of both SC and FC in frontal, parietal and temporal lobes were consistent with results obtained at the whole-brain level. Occipital lobes showed a different profile, with a global FA reduction in AgCC, but similar streamlines count and FC for intra-hemispheric and inter-hemispheric connectivity compared to the TDC group. This reduction in FA values observed for all intra-hemispheric and inter- hemispheric occipital streamlines could reflect sparse, poorly myelinated or divergent fibres (39, 40). Regarding inter-hemispheric streamlines, these results are consistent with the potential existence of an atypical pathway connecting primary visual areas via the anterior commissure in individuals with AgCC showing a reduction in mean FA (11, 12). The reduction of mean FA in intra-hemispheric streamlines in occipital lobes could be associated with the presence of colpocephaly, a congenital brain abnormality characterised by enlargement of the posterior or rear portion of the lateral ventricles, almost uniformly seen when the corpus callosum is absent. This typically results in an enlargement of occipital horns of the lateral ventricles and is known to affect the development of occipital lobes, including white matter integrity in these regions (19, 41, 42).

Different patterns of association between SC and neurobehavioural measures were observed between the AgCC and TDC groups. While in TDC, increased inter-hemispheric SC were associated with better neurobehavioural outcomes (including verbal learning and memory, as well as attentional and executive functioning), we observed the reverse pattern in AgCC with higher intra-hemispheric SC and reduced inter-hemispheric SC associated with these neurobehavioural domains. The pattern observed in the AgCC group is in line with the young age plasticity privilege or “Kennard principle” (3, 43). A very early disruption of embryological callosal development might lead to a complete (or almost complete) absence of axons crossing the midline. If disruption occurs slightly later in gestation, an increased number of callosal fibres might cross the midline and form inter-hemispheric tracks (6, 44, 45). Therefore, it seems that a greater neuroplastic response and brain reorganisation arise in the case of a marked reduction in inter-hemispheric pathways, observed when callosal disruption occurs at an earlier embryological stage. Conversely, and consistent with the literature, TDC seem to rely mostly on inter-hemispheric SC for verbal learning and memory, attentional and executive functioning such as processing speed, switching and verbal fluency, as well as general cognitive abilities (46–50).

Patterns of association between FC and neurobehavioural outcomes were comparable for children with AgCC and TDC. Both intra-hemispheric and inter-hemispheric FC were positively associated with executive and attentional functioning, including visual scanning, selective attention and processing speed. Remarkably, dual task measures (including multi- or uni-modal divided attention and working memory), showed a comparable positive association with increased intra-hemispheric and reduced inter-hemispheric SC in the two groups. However, FC also revealed distinct patterns of association between the two groups. In the AgCC group, higher dual task scores were associated with an overall increase in FC, whereas higher scores in the TDC group were associated with a global decrease in FC. These phenomena could be linked to hemispheric symmetry and parallel processing during divided attention task, as previously observed during dichotic listening in individuals with AgCC (51). However, this requires further investigation. Together, these findings show that the neuroplastic responses observed at the structural level might, at least partly, explain the heterogeneity in neurobehavioural outcomes in the AgCC population.

A major strength of our study is the use of both functional and structural connectivity with a comprehensive set of neurobehavioural measures in a large cohort of children with AgCC. To our knowledge, the present study is the first to use a whole-brain data-driven approach in this population. In light of the literature, our cohort is uniquely large for this pathology. Studies combining neuroimaging and behavioural assessment in this population are rare, and sample sizes are typically smaller than in the present study (11, 52, 53). Nevertheless, our findings should be considered in light of some limitations. Our cohort is heterogeneous in terms of corpus callosum agenesis (i.e., complete or partial AgCC) and associated brain malformations (i.e., isolated or complex AgCC). Further investigations with larger samples of each phenotypic group, possibly achieved through recruitment across multiple sites, are necessary to replicate our findings. Such an approach would also allow exploring the heterogeneous clinical and neuroimaging profiles of children with AgCC, and the factors that might contribute to a better understanding of neurobehavioural outcomes. The combination and extension of these findings in animal models could also significantly improve our understanding of the mechanisms involved. Finally, future studies using graph analysis of structural and functional neuroimaging data combined with clinical measures could lead to a better characterisation of brain dysfunction in terms of aberrant reconfiguration of brain networks in AgCC.

## Conclusions

In conclusion, we provide evidence of structural strengthening of intra-hemispheric pathways in children born without corpus callosum, which seems to allow for functional connectivity comparable to a typically developing brain. In children with AgCC, this increase in intrahemispheric SC, probably reflecting very early disruption of callosal development during gestation, seems to be associated with better verbal learning and long-term memory, verbal working memory, as well as executive and attentional functioning. This work provides novel evidence improving our current understanding of the properties of structural and functional neuroplastic responses in individuals with AgCC. In turn, these neuroplastic responses might be relevant to explain the broad range of clinical presentations and the high heterogeneity in neurobehavioural outcomes observed in this population. Our findings may apply more broadly to other congenital brain malformations in which a similar process of compensation by in utero reorganisation may occur, as well as to neurodevelopmental disorder in which corpus callosum alteration is observed such as autism spectrum disorder, attention deficit hyperactivity disorder or developmental delay.

## Data Availability

Ethical restrictions prevent us from making anonymized data available in a public repository. There are restrictions on data related to identifying participant information and appropriate ethical approval is required prior to release. Only de-identified data will be available (contact).

## ACKNOWLEDGMENTS AND DISCLOSURES

This study was supported by the Boninchi Foundation from the University of Geneva; the Victorian Government’s Operational Infrastructure Support Program; and the Murdoch Children’s Research Institute. Professor Amanda Wood is supported by a European Research Council Consolidator Fellowship [682734]. Associate Professor Richard Leventer is supported by a Melbourne Children’s Clinician Scientist Fellowship. Professor Vicki Anderson was supported by the Australian National Health and Medical Research Council Senior Practitioner Fellowship.

We gratefully acknowledge the families who participated in this study and Kate Pope for her assistance in recruitment of the families.

All authors report no biomedical financial interests or potential conflicts of interest.

## Notes

### Competing Interest Statement

The authors have declared no competing interest.

### Author Declarations

This project was approved by the Royal Children s Hospital (RCH) Human Ethics in Research Committee.

